# Altered herpesvirus-specific immune response is associated with long COVID

**DOI:** 10.64898/2026.11.11.26367525

**Authors:** Ruchika V Joshi, Rizwan MR Khan, Katherine JL Jackson, Sara Ballouz, Brendan Jacka, Leon Ni, Nicholas Findell-Aghnatios, Anupriya Aggarwal, Stuart G Turville, Manu Singh, Cynthia Yau, Ki Wook Kim, Anthony Byrne, Bruce J Brew, Gregory J Dore, David R Darley, Gail V Matthews, Anthony D Kelleher, Chansavath Phetsouphanh

## Abstract

Long COVID is associated with persistent immune dysregulation, but whether immunity to latent viruses is also altered remains unclear. We profiled herpesvirus-specific antibodies and T-cell responses in people with long COVID and recovered convalescent controls at 4, 8 and 24 months after SAR-CoV-2 infection. Overall antibody reactivity and abundance against cytomegalovirus (CMV) and Epstein-Barr virus (EBV), responder frequencies and circulating B-cell subsets were broadly preserved, but early antibody avidity and virus-specific CD4⁺ response magnitude were reduced. Neither virus’s nucleic acid was detected in plasma or stool at 4 or 8 months. Here, we show that long COVID is associated with selective, time-dependent alteration of herpesvirus-specific immune memory, including relatively preserved or enhanced CD8⁺ responses and greater representation of selected herpesvirus-associated T-cell receptor clonotypes. Together, these findings indicate altered herpesvirus-specific antibody quality despite broadly preserved antibody abundance. Lower EBV avidity was associated with poorer quality of life and unrecovered status.

## INTRODUCTION

Long COVID (LC), or Post-Acute Sequelae of COVID-19, can develop following SARS-CoV-2 infection and is associated with symptoms that persist beyond the acute illness. The World Health Organisation estimates that approximately 6% of people with COVID-19 develop LC, although prevalence estimates vary with case definition, follow-up, vaccination status and circulating variants ^1^. A recent report estimates that LC may affect ∼400 million people worldwide and impose an annual economic cost of ∼US$1 trillion, equivalent to approximately 1% of global GDP, although estimates remain highly heterogeneous^2^. This burden highlights that LC is not a transient consequence of COVID-19, but a major and ongoing global health, societal and economic challenge.

Currently, the biological basis of LC remains incompletely understood. In our prospective ADAPT cohort, we previously reported persistent systemic inflammation and altered innate and T cell phenotypes at 8 months after infection^3^. In subsequent follow-up of the same cohort, differences in several measured immune parameters were no longer evident at 24 months, while health-related quality of life had improved for most participants^4^. These findings indicate that immune and clinical features can change over time, but the biological features associated with ongoing symptoms and poorer health status remain unclear.

Reactivation of Herpes viruses, Epstein-Barr virus (EBV) and cytomegalovirus (CMV), which establish lifelong infection, has been proposed as a potential driver of LC. Several studies have associated EBV early-antigen, viral DNA or elevated EBV antibody titres with post-COVID symptoms, although the findings vary across cohorts and sampling periods^5,6^. A recent longitudinal study in people hospitalised with acute COVID-19 documented reactivation of several persistent viruses, including EBV and CMV, with distinct temporal patterns^7^. These results establish that viral reactivation can occur during acute infection, but do not assess herpesvirus-specific immunity during convalescence or its association with persistent symptoms and health status.

Here, we compared EBV- and CMV-specific humoral and cellular immunity in participants with LC and recovered convalescent controls (CC) across follow-up at 4, 8 and 24 months. We assessed seropositivity, antibody abundance and avidity, virus-specific CD4⁺ and CD8⁺ T cell responses, and antigen-associated T cell receptor clonotypes. We also examined associations between EBV antibody avidity and EQ-5D-5L health status and EQ visual analogue scale scores, and measured EBV and CMV nucleic acid in stool swab and plasma. This enabled assessment of whether herpesvirus-specific immune measures were associated with LC and clinical status over time, while distinguishing these associations from evidence of circulating viral nucleic acid.

## Results

### Long COVID is associated with distinct herpesvirus antibody reactivity

To determine whether long COVID (LC) was associated with changes in antiviral humoral immunity, we profiled antibody reactivity to various viral antigens using the Serimmune SERA bacterial-display platform at 4, 8 and 24 months after infection and compared to recovered convalescent controls (CC) (Fig. S1). All samples tested showed no seropositivity for Dengue, Hep C, HIV-1/2 or HTLV-1/2. Overall seropositivity in LC and CC showed broadly similar reactivity across the herpesvirus panel (CMV, HSV-1, HSV-2, EBV lytic and EBV latent (Fig. S1)), with no evidence of a generalized loss of herpesvirus recognition over time, although seropositivity of LC to EBV late antigens was higher than CC (80% and 61%, respectively; *p=* 0.043) (Fig. 1a). Continuous antibody-reactivity scores revealed that CMV and EBV lytic, latent and late-antigen responses were heterogeneous, with substantial inter-individual variation and no consistent directional shift across all visits (Fig. 1b).

**Fig 1:**
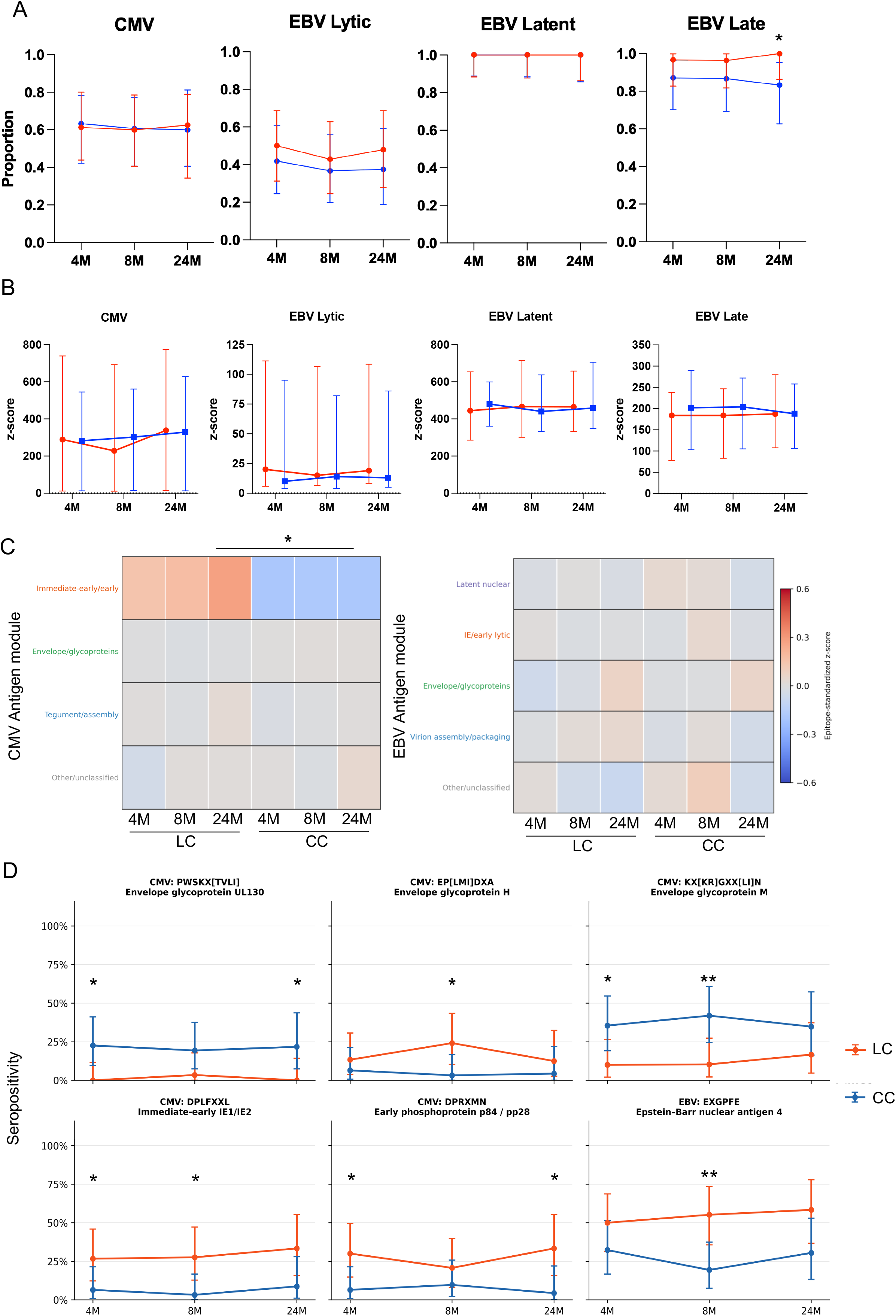
Long COVID is associated with altered herpesvirus antibody reactivity. **a,** Seropositivity to CMV, and EBV lytic, latent and late proteins at 4, 8 and 24 months in individuals with long COVID (LC) and matched controls (MC), measured using the Serimmune bacterial-display platform. **b,** Longitudinal abundance of herpesvirus-specific antibody reactivity, shown as participant-level z-scores across follow-up. **c,** Heatmaps showing module-level antibody reactivity for CMV and EBV antigen groups. **d,** Longitudinal seropositivity for individual CMV and EBV epitopes that differed nominally between LC and MC by Fisher’s exact test. Symbols and error bars indicate group summaries and variability. FU4, FU5 and FU8 correspond to 4-, 8- and 24-month follow-up, respectively. LC, red; MC, blue; *p* values <0.05 (*), <0.01 (**), <0.001 (***) and <0.0001 (****) were considered significant.

We next examined whether these differences were distributed across antigenic protein module/groups. CMV module analysis identified a selective difference in immediate-early/early antigen reactivity, which was higher in LC at 24 months (LC median standardised z-score 0.42 versus CC median −0.123; *p=* 0.013)(Fig. 1c). The remaining CMV modules, including envelope/glycoprotein and tegument/assembly antigens, showed limited separation between groups. EBV group-level responses were comparatively conserved, with latent, lytic/early, envelope and virion assembly/packaging modules showing overlapping patterns between LC and CC across 24 months. At greater resolution, individual epitope analysis identified modest differences in seropositivity between LC and CC (Fig. S2). For CMV, LC participants showed lower seropositivity to an UL130 envelope glycoprotein epitope at 4 months (LC 0% versus MC 22.6%; *p=* 0.011) and 24 months (*p=* 0.021), and to a glycoprotein M epitope at 4 (*p=* 0.031) and 8 months (*p=* 0.008)(Fig. 1d). In contrast, seropositivity to an IE1/IE2-associated epitope and to the p84/pp28-associated epitope DPRXMN was higher in LC at 4 and 24 months (*p=* 0.043 and *p=* 0.022, respectively). An additional difference was observed for a glycoprotein H epitope at 8 months ((Fig. 2f, 0.024). EBV differences were more restricted, where seropositivity to the EBNA4 epitope EXGPFE was higher in LC than MC at 8 months (LC 55.2% versus MC 19.3%; *p=* 0.007), whereas other EBV epitopes showed broadly overlapping longitudinal profiles.

**Fig. 2:**
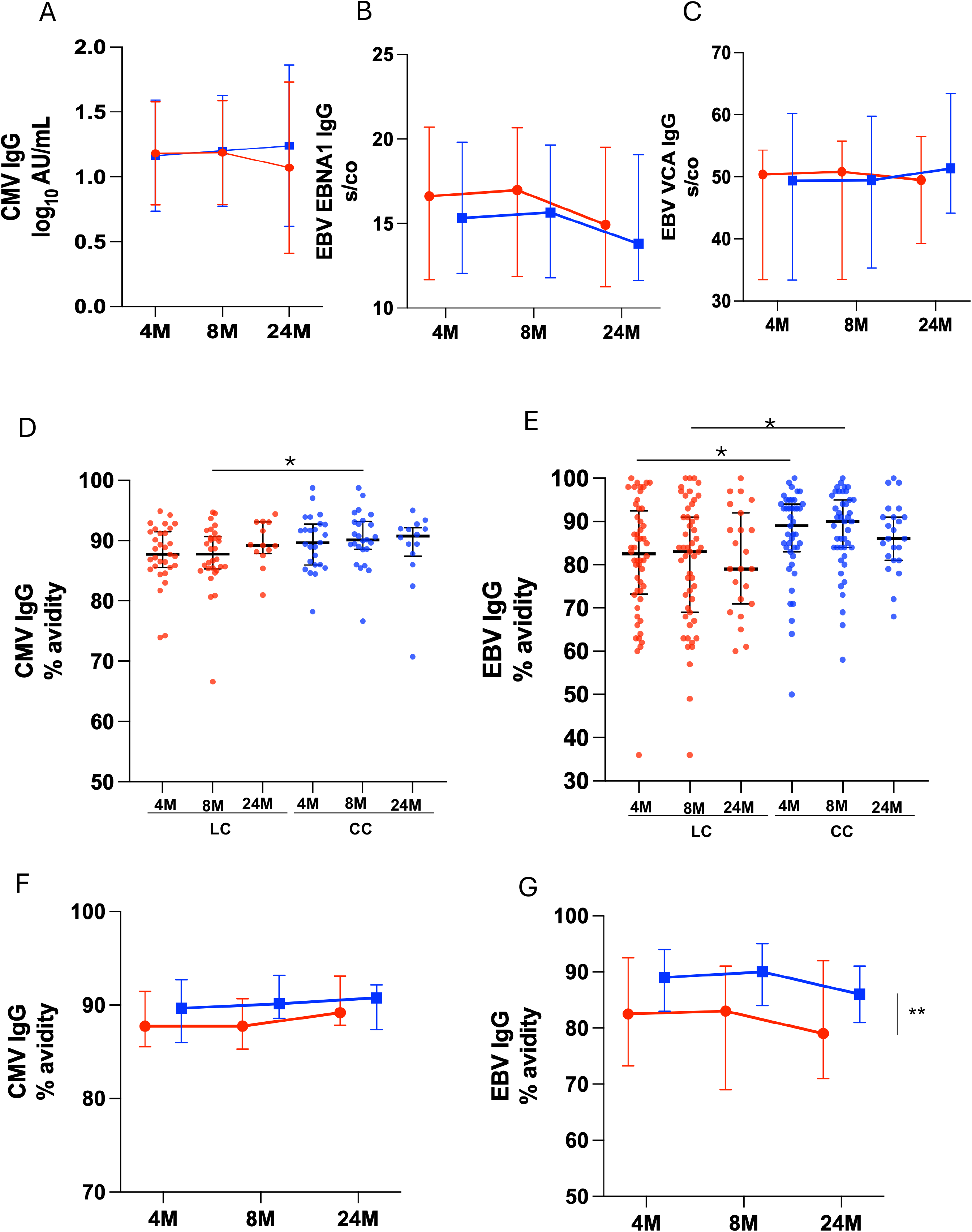
Reduced CMV and EBV antibody avidity despite preserved total IgG. **a**, Longitudinal concentrations of total CMV IgG, EBV EBNA1 IgG (**b**) and EBV VCA IgG (**c**) at 4, 8 and 24 months. CMV IgG is shown as log10 AU ml⁻¹, and EBV IgG measurements are shown as signal-to-cut-off ratios. **d, e,** CMV and EBV IgG avidity across follow-up. **f,** Longitudinal group summaries of CMV and EBV avidity in LC and MC. Individual points represent participants; central lines and error bars indicate group-level estimates and variability. Comparisons were performed using mixed-effects models and prespecified post hoc tests. LC, red; MC, blue; *p* values <0.05 (*), <0.01 (**), <0.001 (***) and <0.0001 (****) were considered significant.

### Reduced CMV and EBV antibody avidity despite preserved total IgG

CMV and EBV serology was performed on Abbott Alinity instrument, total CMV IgG and EBV-specific IgG concentrations were broadly comparable between individuals with long COVID (LC) and matched controls (MC) across follow-up. This included CMV IgG, EBV EBNA1 IgG and EBV VCA IgG, with no clear separation between groups at 4, 8 or 24 months (Fig. 2a, b, c). In contrast, antibody avidity was reduced in LC. CMV IgG avidity was lower in LC than CC at the early follow-up visits, with significant differences at month 8 (median LC 66.6% (IQR, 28.07-94.68%); MC, 76.66% (IQR, 22.06-98.72%, *p=* 0.044) whereas group distributions were more similar by FU8 (Fig. 2d). EBV IgG avidity showed a comparable trend, with lower avidity among LC participants at 4M and 8M and partial convergence at 24M (Fig. 2e). Notably, two LC participants had positive IgM levels to CMV and EBV (data not shown). To determine temporal changes, mixed-effects analysis was performed, no statistically significant effect was observed for CMV avidity (Fig. 2f), but an overall effect of clinical group on EBV IgG avidity was evident across follow up. Model-estimated avidity showed significant effect on clinical group over time, where avidity was lower in LC than in MC (80.65% versus 87.40%; estimated difference, −6.75 percentage points; 95% CI, −11.51 to −1.99; *F*(1,93) = 7.94, *p=* 0.0059) (Fig. 2g). There was no significant effect of time (*F*(1.81,123.3) = 0.035, *p=* 0.955) or time-by-group interaction (*F*(1.81,123.3) = 1.11, *p=* 0.327).

### CMV and EBV nucleic acid was not detected in plasma or stool swabs

Following the observed differences in CMV- and EBV-specific antibody avidity, we assessed whether viral nucleic acid was detectable in plasma or stool swab specimens collected at 4 and 8 months. Virome capture sequencing identified several viral taxa in the stool swabs, but neither CMV nor EBV met the threshold of at least 100 species-level BLAST-matched reads in more than three samples (Fig. S3). Consistent with this, CMV and EBV DNA was not detected by plasma qPCR in any LC or CC samples (data not shown). Taken together, although herpesvirus-specific antibody avidity differed between groups, we found no evidence of detectable CMV or EBV nucleic acid in the sampled plasma or stool specimens at these visits.

### Reduced virus-specific CD4⁺ and enhanced CD8⁺ T-cell responses in long COVID

Circulating CD19+ B-cell subset distributions were broadly comparable between individuals with LC and CC at 4 and 8 months. The proportions of naïve (IgD+CD27-), atypical (double-negative; IgD-CD27-), non-switched memory (IgD+CD27+) and switched-memory B cells (IgD-CD27+) were largely preserved, with no consistent evidence of general B-cell compartment remodelling (Fig. 3a). Although atypical subset was higher in CC compared to LC at 8M (*p=* 0.015), this did not represent a consistent pattern across B-cell subsets.

**Fig. 3:**
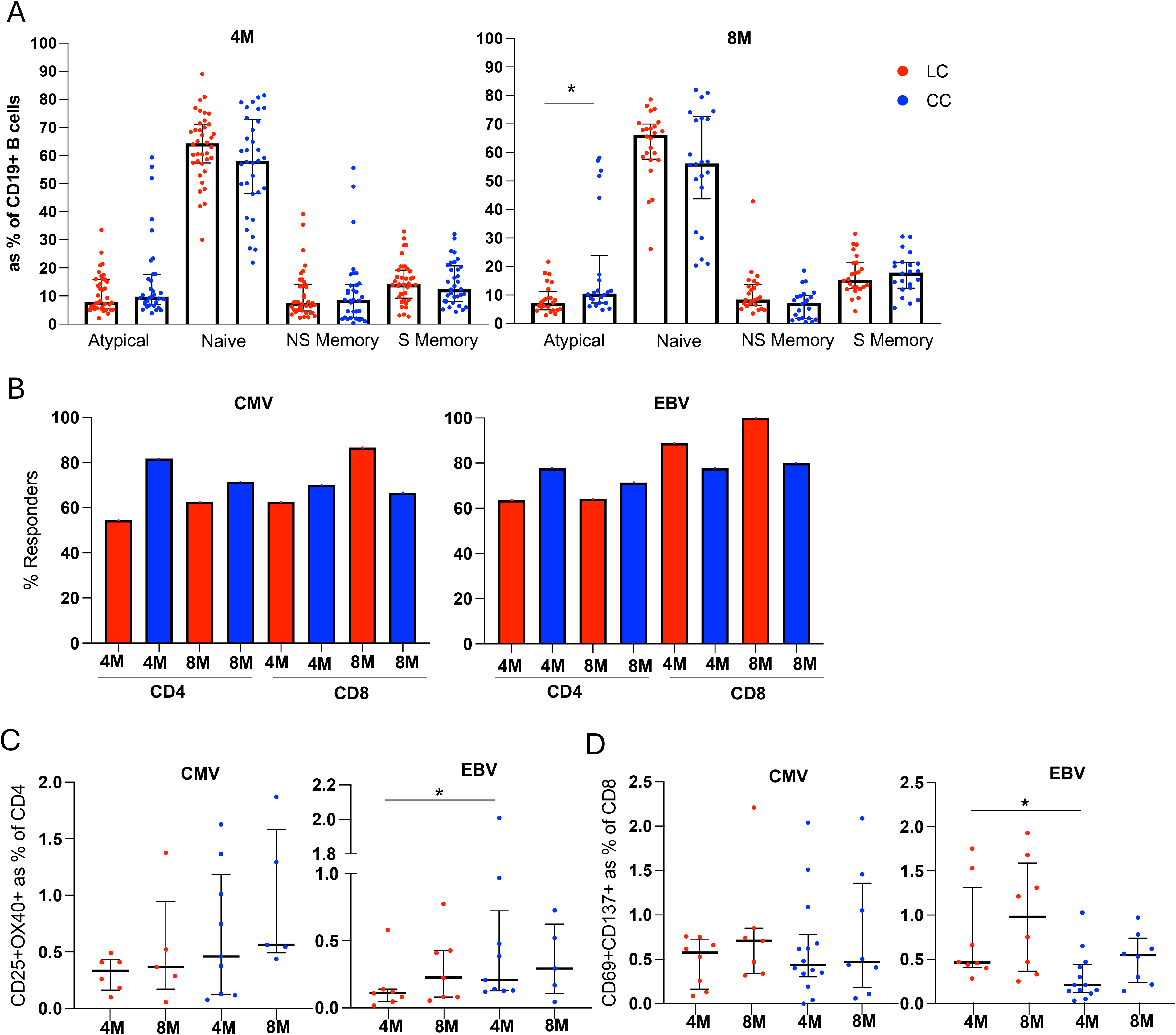
Reduced virus-specific CD4⁺ but preserved or increased CD8⁺ T-cell responses in long COVID. **a,** Frequencies of circulating CD19+ B-cell subsets at 4 months (4M) and 8 months (8M), including Atypical (double-negative; IgD-CD27-), naïve (IgD+CD27-), non-switched memory (IgD+CD27+) and switched-memory (IgD-CD27+) B cells. Data are shown as a percentage of the parent B-cell population. **b,** Proportion of LC and MC participants with detectable CMV- or EBV-specific CD4⁺ and CD8⁺ T-cell responses at 4 and 8 months. **c,** Magnitude of CMV pp65- and EBV-specific CD4⁺ T-cell responses measured by the activation-induced marker assay. CD4⁺ responses were defined by CD25 and OX40 co-expression. **d,** Magnitude of CMV pp65- and EBV-specific CD8⁺ T-cell responses, defined by CD69 and CD137 co-expression. Each point represents an individual participant; bars indicate group summaries and variability. LC, red; MC, blue. Comparisons were performed using two-sided non-parametric tests, *p* values <0.05 (*), <0.01 (**), <0.001 (***) and <0.0001 (****) were considered significant.

Activation induced marker (AIM) assays showed a distinct pattern of virus-specific T-cell responses. The proportion of participants mounting detectable CMV- or EBV-specific CD4⁺ or CD8⁺ responses were broadly similar between LC and CC at the assessed timepoints (Fig. 3b). However, analysis of response magnitude revealed reduced virus-specific CD4⁺ responses in LC. CMV pp65-specific CD4⁺ responses were lower in LC than CC at 4 and 8 months, although these differences did not reach statistical significance. EBV-specific CD4⁺ responses were significantly reduced in LC at 4M (*p=* 0.024), whereas the groups were similar at 8M (Fig. 3c). In contrast, CD8⁺ T-cell responses were maintained or increased in LC. CMV pp65-specific CD8⁺ responses did not differ significantly between LC and CC at either timepoint (Fig. 3d). EBV-specific CD8⁺ responses were higher in LC at month 4 (*p*= 0.0133), with the difference lessened by month 8.

### Persistent expansion of CMV- and EBV-associated T-cell clonotypes in long COVID

Single-cell gene expression (GEX) and T cell receptor (TCR) sequencing from n=13 CC and n=32 LC identified a diverse repertoire of antigen-associated clonotypes across follow-up. Both GEX and TCR V(D)J data were obtained for 29,218 cells from CC and 74,659 cells from LC. Clustering based on GEX data identified T cells subsets (Fig 4a) and permitted the projection of clonotypes annotated with CMV, EBV, SARS-CoV-2 and other viral and microbial specificities across the T cell landscape (Fig. 4b). SARS-CoV-2 annotated clonotypes were the most common for both CC (1.44% of clonotypes) and LC (1.38% clonotypes), and CMV- and EBV-associated clonotypes were detected in both LC (CMV: 0.675%, EBV: 0.182%) and CC (CMV: 0.386%, EBV 0.108%). For LC, CMV clonotype were prominently persisting clonotypes observed at both the 4M/8M (p=2.15e-36; Fisher Exact with BH adjustment), or 4M, 8M and 24M (*p*=0.0284; Fisher Exact with BH correction). EBV clonotypes were more frequent in LC than CC at 4M (*p* < 0.0001), 8M (*p* < 0.0001) and 24M (*p*=0.0182) but LC EBV clonotypes were not significantly enriched for clonotypes detected across multiple time points (Fig. 4c).

**Figure 4.**
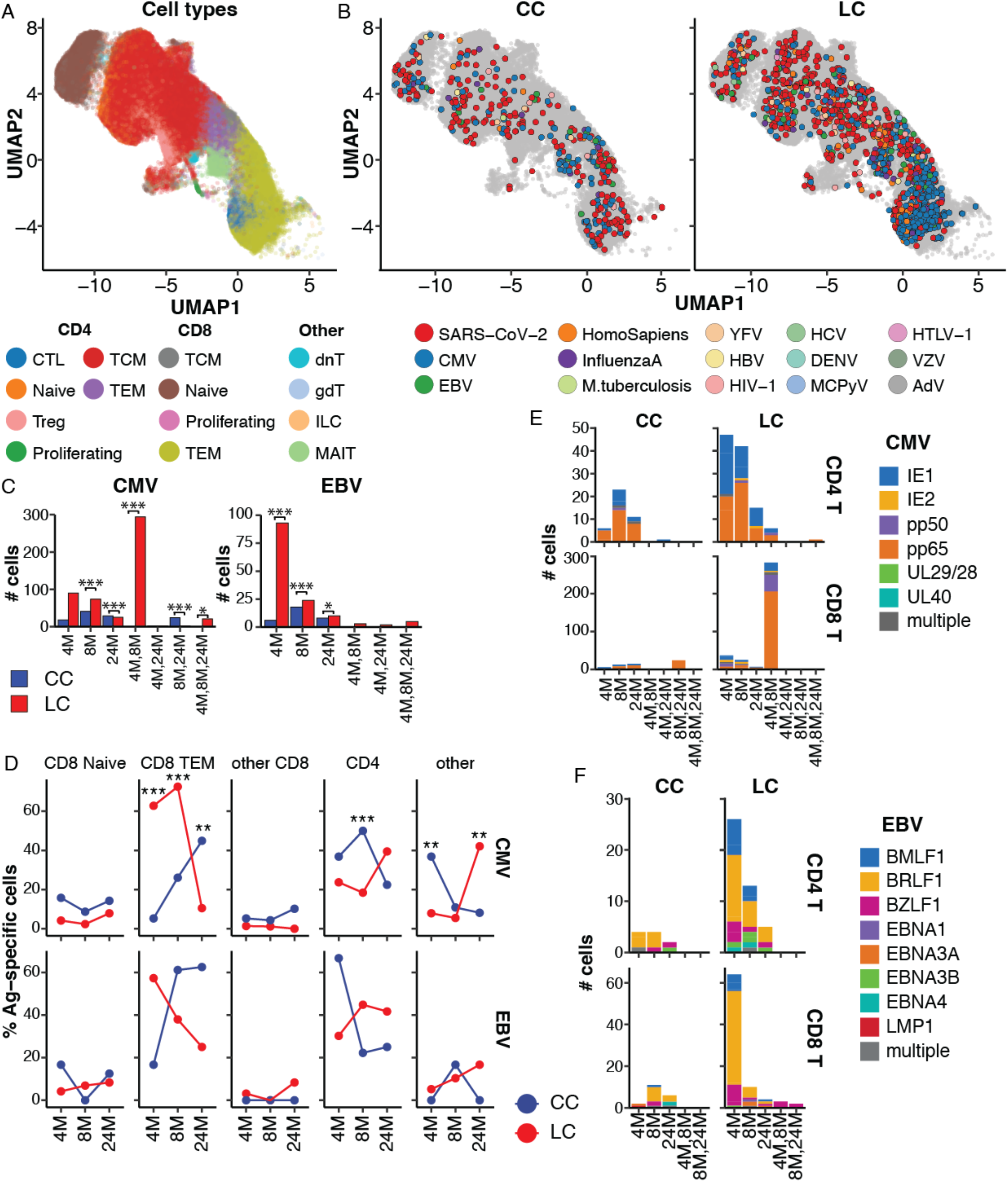
Long covid is associated with increased T cells with CMV- and EBV- associated TCRs. **a,** UMAP of T cell subsets from single cell gene expression analysis for T cells with matched T cell V(D)J sequencing for n=13 CC and n=32 LC donors. Cell annotations from Azimuth. **b,** UMAP projection with cells coloured for clonotype annotation from comparison to TCRs of known antigen specificity (exact CDR3 matched, same TRBV and TRBJ usage). Cells without annotation or annotated for >1 antigen are coloured in grey. **c,** Count of CMV and EBV annotated cells for LC and CC based on clonotype distribution; single time point (4M, 8M or 24M) or clonotypes that persisted across >1 follow-up. **d,** T cell subset distribution of CMV (upper) or EBV (lower) annotated clonotypes, data points indicate the proportion of Ag-annotated cells from LC or CC that were observed a given T cell population at each follow-up. **e,** Mapping of CMV specific clones to proteins by clonotype follow-up occurrence. **f,** Mapping of EBV specific clones to proteins by clonotype follow-up occurrence.

Comparison of annotated clonotypes within T cell subsets showed differences in LC and CC among T cell subsets for CMV (Fig. 4d), with an increased proportion of CD8 TEM for LC compared to CC at 4M (62.8% vs 5.26%, *p* < 0.0001) and 8M (72.5% vs 26.1%, *p* < 0.0001) follow-up. EBV did not show any proportional differences within the different CD4 and CD8 subsets across the follow-ups (Fig 4d, all p-values > 0.05). Epitope mapping within EBV and CMV annotated clonotypes showed that CMV- reactive clonotypes targeted multiple viral proteins, including IE1, IE2, pp50, pp65 and UL29/28 (Fig 4e). EBV-associated clonotypes similarly included specificities for BMLF1, BZLF1, EBNA3A and EBNA4 (Fig. 4f). Thus, the expanded repertoire was distributed across several viral antigens rather than being dominated by a single protein.

### Lower herpesvirus antibody avidity is associated with impaired quality of life measures

To examine the clinical relevance of altered antibody avidity, CMV- and EBV-specific antibody measures were integrated with clinical, biological and quality-of-life variables. At month 4, EBV avidity correlated positively with EBV VCA IgG (Spearman’s ρ= 0.52, *p*< 0.001), EBV EBNA1 IgG (ρ= 0.34, *p=* 0.016), SARS-CoV-2 spike IgG (ρ= 0.30, *p=* 0.030) and BMI (ρ= 0.42, 0.004). EBV avidity was inversely correlated with CMV avidity (ρ= −0.36, *p=* 0.050). At 8 months, EBV avidity correlated positively with EBNA1 IgG (*ρ* = 0.286, *p=* 0.042) and VCA IgG (*ρ*= 0.484, *p*< 0.001). Positive correlations were also observed with breathlessness (*ρ*= 0.333, *p=* 0.002) and VAS fatigue (*ρ* = 0.252, *p=* 0.032). Avidity did not correlate with inflammatory measures, including CRP, D-dimer or lymphocyte counts. Integrated Principal component analysis showed greater spread of LC participants than controls at both follow-up visits, but with substantial overlap between groups (Fig. 5b). At 4 months, PC1 and PC2 explained 20% and 8% of the variance, respectively. EQ-index and EQ-VAS loaded negatively on PC1 (−0.33 and −0.31), whereas phantosmia, troponin and anosmia loaded positively on PC2 (0.35, 0.35 and 0.33); EBV avidity also loaded positively on PC2 (0.29), while D- dimer loaded negatively (−0.29). Twelve LC participants with low EBV avidity occupied a visually distinct, dispersed region of the PCA. This pattern was not observed at 8 months.

**Fig. 5:**
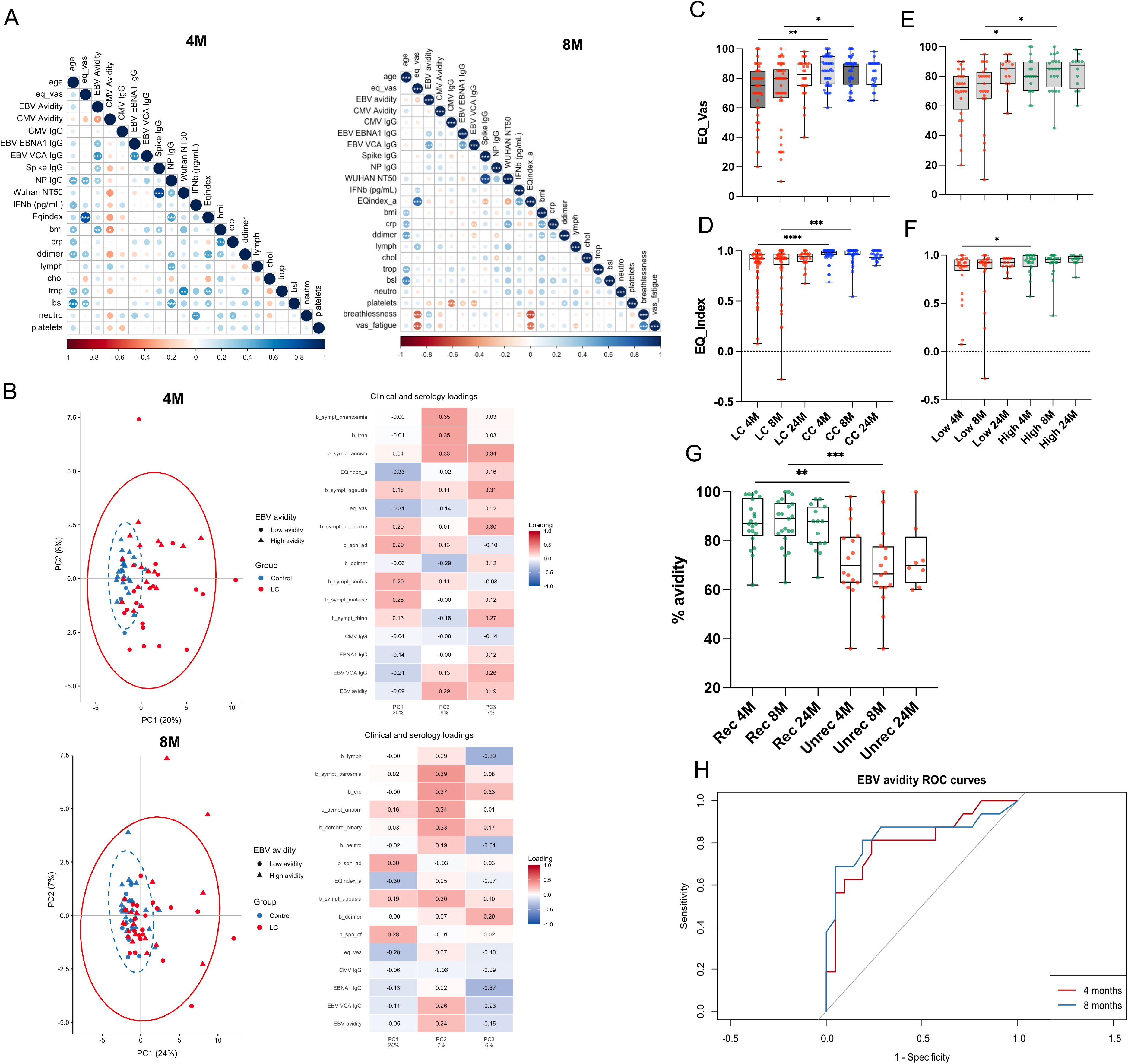
Clinical associations of herpesvirus antibody avidity. **a,** Spearman correlation matrices integrating clinical, serological and inflammatory variables at 4 and 8 months. Variables included CMV and EBV antibody avidity, CMV and EBV IgG concentrations, SARS-CoV-2 spike and nucleoprotein IgG, Wuhan neutralisation titres, interferon-β, EQ-VAS, EQ-index, body mass index, CRP, D-dimer, lymphocyte count and other clinical laboratory measures. Colour indicates the direction of correlation and circle size indicates its magnitude. **b,** Principal component analyses integrating clinical and serological variables at 4 and 8 months. Points are coloured by clinical group and shaped according to low- or high-avidity status. Loading plots show the contribution of individual clinical and serological variables to the principal components. **c, d,** EQ-VAS and EQ- index, respectively, in LC and CC at 4, 8 and 24 months. **e, f,** EQ-VAS and EQ-index, respectively, in LC participants stratified by EBV avidity at each visit using the exploratory 83% threshold. **g,** EBV avidity over time in participants classified as recovered or unrecovered using an EQ-index threshold of 0.9 at later follow-up. **h,** Receiver operating characteristic (ROC) curves for EBV avidity at 4 and 8 months to distinguish recovery groups. AUCs were 0.808 at 4 months and 0.839 at 8 months; the AUCs did not differ significantly (*p* = 0.505). In **c–g**, boxplots show the median and interquartile range, with individual participants overlaid. Recovery was defined as EQ-index >0.9. Asterisks indicate statistical significance, *p* values <0.05 (*), <0.01 (**), <0.001 (***) and <0.0001 (****).

LC participants had lower EQ-VAS than CC at 4 and 8 months (*p=* 0.002 and *p=* 0.039, respectively), with no difference at 24 months (Fig. 5c). EQ-index was also lower in LC at 4 and 8 months (*p*< 0.0001 and *p*< 0.001, respectively), but not at 24 months (Fig. 5d). Within LC, participants with EBV avidity below the exploratory 83% threshold generally had lower EQ-VAS at 4 months (median 72.5, versus 80.0, *p=* 0.024) and 8 months (75.0, versus 85.0, *p=* 0.014)(Fig. 5e), than those above the threshold; EQ-index showed a similar pattern, evident at 4 months (median 0.890, versus 0.9505, *p=* 0.026), but not at 8 months (Fig. 5f). These findings suggest that the threshold may help identify a subgroup with poorer reported health status.

When participants were classified by recovery status at later time points (18-24months) using an EQ- index threshold of 0.9, unrecovered LC participants had lower EBV avidity than recovered controls longitudinally, with the clearest group contrasts at 4 (median 70, versus 82, *p=* 0.005) and 8 months (median 66.5, versus 89, *p=* 0.002) (Fig. 5g). We next used receiver operating characteristic (ROC) analysis as recovery status was binary and EBV avidity was measured continuously. The area under the curve (AUC) therefore quantifies discrimination, rather than overall classification accuracy at a single threshold. ROC analyses showed discrimination between these recovery groups by EBV avidity at 4 months (AUC = 0.808, *p=* 0.002) and 8 months (AUC = 0.839, 95% CI 0.691-0.988, *p*< 0.001).

The AUCs for the two timepoints did not differ significantly (*p=* 0.505), so the numerically higher estimate at 8 months does not demonstrate better discrimination than at 4 months. At 24 months, the AUC was 0.783 (*p=* 0.028), based on 15 recovered controls and 8 unrecovered LC participants, with missing data due to lost to follow up (Fig. 5h and Fig. S4). Together, these exploratory analyses associate lower EBV avidity with poorer health-related quality of life and recovery status.

## DISCUSSION

Our findings identify selective, temporal alteration of herpesvirus-specific immunity in long COVID. Total CMV- and EBV-specific IgG concentrations and the proportions of participants with detectable virus-specific T-cell responses were broadly preserved. In contrast, antibody avidity was lower in LC at 4 and 8 months post-infection, virus-specific CD4⁺ AIM responses were reduced, and selected CMV- and EBV-associated T-cell clonotypes were more prominently represented. Some differences attenuated by 24 months, consistent with partial recovery rather than a uniform or irreversible loss of antiviral immunity. These data extend our previous observations of persistent, but improving, immune dysregulation in the ADAPT cohort by resolving changes in the quality and cellular distribution of antiviral memory^3,4^.

Lower CMV and EBV avidity despite broadly comparable IgG concentrations suggests that antibody quantity alone does not capture the altered humoral response in LC. Reduced avidity is consistent with differences in the maturation or maintenance of virus-specific antibody responses, but our data cannot determine whether this reflects altered germinal-centre selection, changes in memory B-cell recruitment, differences in antigen exposure, or a combination of these processes. Nor does lower avidity establish active herpesvirus replication, as we did not detect CMV or EBV nucleic acid in plasma or stool swabs. These negative findings limit evidence for detectable viraemia or shedding at 4 and 8 months, but do not exclude intermittent or tissue-restricted viral activity below the sensitivity of the assays. This distinction is relevant to prior work by Peluso and colleagues, who reported associations between serological markers suggestive of EBV reactivation and symptom clusters, but no association with ongoing EBV viraemia; CMV seropositivity was associated with lower odds of neurocognitive symptoms in their cohort ^8^. Our results complement that work by examining antibody avidity and antigen-specific cellular responses longitudinally, rather than relying only on seropositivity or antibody titre.

Cellular findings point to a selective shift in the balance of virus-specific T-cell responses. The proportion of participants with detectable CMV- or EBV-reactive AIM responses was similar between groups, whereas response magnitude showed weaker CD4⁺ responses in LC and preserved or increased CD8⁺ responses, including higher EBV-specific CD8⁺ responses at 4 months. Circulating B-cell subset distributions were largely comparable, providing little evidence for broad B-cell compartment remodelling. One possibility is that reduced virus-specific CD4⁺ activity reflects altered helper or coordinated antiviral function, while CD8⁺ responses remain maintained or expand in response to prior or continuing antigenic stimulation. The association between reduced antibody avidity and altered CD4⁺ responses are compatible with a disturbance in the coordination of cellular and humoral memory but does not establish that one causes the other. It also remains possible that the measured response magnitude reflects differences in the composition or functional state of antigen-specific T cells rather than a general defect in antiviral immunity.

The greater representation of selected CMV- and EBV-associated clonotypes in LC adds a repertoire-level dimension to this pattern. Their detection across multiple viral antigens, together with lower CD4⁺ responses and preserved or enhanced CD8⁺ responses, is consistent with persistent remodelling of latent-virus-associated T-cell memory^9,10^. One possible context for this later remodelling is acute viral activity during SARS-CoV-2 infection. Maguire *et al.* detected transcripts from EBV, CMV and other persistent viruses during acute COVID-19 and reported that persistent Anelloviridae detection during convalescence was associated with PASC symptoms^7^. These observations raise the possibility that acute infection-associated viral activity and inflammation could shape long term antiviral memory, even when viral nucleic acid is no longer detectable in plasma or stool at later visits.

The clinical analyses suggest that lower early EBV avidity may mark a subgroup with poorer health-related quality of life and slower recovery. Lower-avidity LC participants had lower EQ-VAS at 4 and 8 months, while EQ-index was lower at 4 months; avidity also discriminated between recovered controls and unrecovered LC participants in exploratory ROC analyses. These results are consistent with previous immune profiling studies identifying heterogeneous biological features associated with LC and symptom burden ^11,5^. However, the avidity threshold was exploratory, the ROC analyses were conducted in the same cohort in which the association was observed, and the AUCs at 4 and 8 months did not differ significantly. The results therefore support further evaluation of EBV avidity as a candidate stratification measure, but do not establish a validated prognostic test or show that avidity itself drives persistent symptoms.

Several limitations should be considered. The cohort size was modest, particularly at 24 months, and missing follow-up data limit precision at this visit. The study sampled plasma and stool swabs rather than potential tissue reservoirs and cannot rule out intermittent or compartmentalised viral activity. The TCR assignments require functional confirmation, and the clinical associations and 83% avidity threshold require independent replication with prespecified analyses and adjustment for relevant covariates and multiple testing. Future longitudinal studies that include acute timepoints, that combine antibody avidity and B-cell lineage (including B-cell receptor sequencing) analysis with functional T-cell assays and viral nucleic acid detection could establish whether altered antibody maturation and clonal expansion share a biological driver, and whether these measures identify reproducible clinical subgroups.

Taken together, our findings support a model in which LC is associated with a persistent, but potentially partially reversible, distortion of herpesvirus-specific immune memory. This phenotype involves altered antibody quality and a modification in the magnitude and clonal representation of virus-specific T-cell responses, without evidence here of detectable CMV or EBV nucleic acid in plasma or stool. Defining the mechanisms and clinical relevance of this immune remodelling will require validation across larger cohorts and direct functional testing.

## METHODS

### Study Design

ADAPT was a prospective observational cohort of adults with confirmed SARS-CoV-2 infection recruited through St Vincent’s Hospital Sydney and affiliated community services during the first wave of the Australian pandemic, mid-2020 to mid-2022. The cohort and longitudinal sampling framework have been described previously^3,12,13^. Participants were followed for up to 24 months after infection, with clinical and biological assessments performed at prespecified visits. Long COVID (LC) was defined by the presence of persistent fatigue or malaise, dyspnoea or chest pain at least 90 days after the estimated date of infection. The study was approved by the St Vincent’s Hospital Sydney Human Research Ethics Committee (2020/ETH00964) and registered with the Australian New Zealand Clinical Trials Registry (ACTRN12620000554965). All participants provided written informed consent before study procedures.

For this immunology substudy 105 participants at enrolment, including 52 participants with long COVID (LC) and 53 convalescent controls (CC) (Supplementary Table S1), were included from the main ADAPT cohort. LC was defined at the 4-month assessment by the presence of at least one of three major symptoms: fatigue, dyspnoea or chest pain. The reported median age was 50.5 years (27– 83) in LC and 50 years (23–75) in CC. The LC group included 29 female (55.8%) and 23 male participants (44.2%); the CC group included 27 female (50.9%) and 26 male participants (49.1%).

White ethnicity was reported for 47 LC participants (90.4%) and 46 controls (86.8%), while 24 LC participants (46.2%) and 25 controls (47.2%) had one or more comorbid conditions. Most LC participants were enrolled from the community (39, 75.0%), whereas nearly all controls were community-enrolled (50, 94.3%). Moderate initial illness was reported in 51 LC participants (98.1%) and 14 controls (26.4%). Participants underwent clinical and immunological assessments at 4, 8 and 24 months after infection.

#### Clinical Measures

##### Long COVID classification

As previously described in the ADAPT cohort^12^, participants were recruited during the initial wave of COVID-19 up to emergence of omicron variant BA.1/2, from hospital and community locations following mild/moderate severity of infection. Long COVID status was assigned if participants reported >1 persistent symptom of dyspnoea, chest pain, or fatigue/malaise at least 90 days after estimated date of initial infection. Age- and sex-matched participants who were asymptomatic were included in the current analysis as matched controls to participants with long COVID.

##### Patient-reported outcomes

Comprehensive patient-reported outcome measures were assessed at all follow-up visits. Generic health-related quality of life was assessed using the EQ-5D-5L tool, which measures five dimensions of health (mobility, self-care, usual activities, pain/discomfort, and anxiety/depression) through a 5-level Likert scale (ranging from no problems to extreme problems). Health state utility values (range –0.25 to 1.00) were calculated for each participant visit based on their responses using the English value set^14^ (version 1.1, updated 01/12/2020). To classify generic “poor health”, participants index scores were compared to age- and sex-matched normative values from the general population of South Australia^15^. Participants were considered to have poor health if their EQ-5D-5L score was below than the lower 95% confidence interval of the population values. It is anticipated that the normative general population values in South Australia are comparable to the normative general population values where the participants were located (i.e., neighbouring state of New South Wales).

To understand how COVID-19 specifically impacts daily activities, participants rated their agreement to four statements about functioning: 1) “I have fully recovered after COVID-19“; 2) “I feel confident returning to my pre-COVID work”; 3) “I have returned to my usual activities of daily living”; and 4) “I have returned to my normal exercise level”. Responses were on a 6-level Likert scale (Strongly Disagree; Disagree; Slightly Disagree; Slightly Agree; Agree; Strongly Agree) and completed at the 24-month visit.

##### Routine pathology

Haematology, biochemistry, and immune biomarkers (C-reactive protein, D-dimer, troponin I, total cholesterol, lymphocyte count, neutrophil count, and blood glucose) were assessed at each study visit as part of routine clinical care. Biomarkers were analysed using NATA accredited clinical chemistry/pathology platforms (SydPATH, St Vincent’s Hospital, Sydney, Australia).

##### CMV and EBV serology

Plasma CMV and EBV serology was performed by Prince of Wales Hospital Diagnostic Services. CMV specific IgG and IgM and EBV specific EBNA IgG, VCA IgG and VCA IgM antibodies were measured using automated serological assays according to the manufacturer’s instructions and laboratory SOP. Results were reported as assay specific index or quantitative values and interpreted according to manufacturers and laboratory validated cut offs. CMV IgG avidity was determined using the Abbott Alinity CMV IgG Avidity chemiluminescent microparticle immunoassay (CMIA)^16^. Briefly, CMV specific IgG binding was assessed using avidity competition format following pretreatment with and without soluble CMV antigen. Percentage IgG avidity is derived from the paired measurements according to the assay formula [100 × (1 − signal following soluble antigen pretreatment/reference signal)] and calculated by the assay software. Higher avidity values indicate a greater proportion of mature, high avidity CMV specific IgG. The EBV IgG avidity was assessed against VCA IgG using a differential dissociation method in parallel with and without 8M urea treatment^17^. EBV VCA-IgG avidity was calculated as the proportion of antibody binding retained following urea treatment relative to the untreated measurement as VCA-IgG avidity (%) = (signal with urea treatment / signal without urea treatment) × 100^18^. Lower avidity values reflected dissociation of recently generated, low avidity VCA-IgG, whereas higher values reflected mature, high avidity VCA-IgG.

#### Laboratory Experiments

##### Bacterial Display Antibody Screen

Serum antibody reactivity was profiled by Serimmune (USA) using its Serum Epitope Repertoire Analysis (SERA) bacterial-display platform. In this assay, antibodies bind peptide mimotopes displayed on a diverse *Escherichia coli* library; antibody-bound bacteria are enriched by magnetic selection, and the peptide-encoding DNA is sequenced to identify and quantify enriched motifs. Enriched motifs are mapped to viral proteomes to infer epitope-level antibody reactivity. Serimmune-provided epitope z-scores were used for continuous analyses. These scores represent motif enrichment standardised relative to the assay’s negative-control reference distribution; epitope reactivity was classified as seropositive when the z-score was >4. CMV and EBV epitopes were assigned to antigen modules according to their annotated viral proteins, independently of their statistical association with clinical group. For each participant and visit, module reactivity was calculated as the arithmetic mean of the available epitope z-scores assigned to that module. The CMV modules comprised immediate-early/early, envelope/glycoprotein, tegument/assembly and other/unclassified antigens. The EBV modules comprised immediate-early/lytic, latent nuclear, envelope/glycoprotein, virion assembly/packaging and other/unclassified antigens.

##### Ex vivo phenotyping and combined CD4/CD8 T cell activation induce marker assay

Cryopreserved PBMCs were thawed using RPMI medium containing L-glutamine and 10% FCS (ThermoFisher Scientific, USA) supplemented with Penicillin/Streptomycin (Sigma-Aldrich, USW) and subsequently stained with monoclonal antibodies (mAb) binding to extracellular markers. Extracellular panel included: Live/Dead dye Near InfraRed, CD38 (HIT2, #MHCD3819 [1:100]) (ThermoFisher Scientific, USA); CD3 (UCHT1, # 300430 [1:100]), CD8 (HIL-72021, # 301042 [1:100]), CD123 (6H6, #306043 [1:50]), PD-1 (EH12.1, #562516 [1:100]), TIM-3 (TD3, #746771[1:100]), CD27 (L128, #563167 [1:100]), CD45RA (HI100, #564552 [1:100]), IgD (IA6-2, #561315 [1:100]), CD25 (2A3, #340939 [1:20]), and CD19 (HIB19, #557921 [1:100]) (BioLegend, USA); CD4 (OKT4, 300533 [1:100]), CD127 (A019D5, #351325 [1:100]), HLA-DR (L234, #307671 [1:100]), CCR7 (G043H7, 353217 [1:100]), CD16 (GB11, #302054 [1:100]), CD14 (HCD14, #325631 [1:100]), CD56 (NCAM-1, #557747 [1:100]), CD11c (B-ly6, #561355 [1:20]), and CD57 (QA17A04, #393304[1:100]) (BD Biosciences, USA). FACS staining of 48hr activated PBMCs was performed as described previously, but with the addition of CD137 (4B4-1) to the cultures at 24hrs. Final concentration of 10µg/mL of SARS-CoV-2 peptide pools^19^, CMV pp65 and EBV 15mer peptide pools (Genscript) were used and staphylococcal enterotoxin B (SEB; 1 µg/ml)(#S4881-MG, Sigma) was used as a positive control (ThermoFisher Scientific). In vitro activation mAb panel included: CD3 (UCHT1, # 300429 [1:100]), CD4 (RPA-T4, # 557922 [1:100]), CD8 (RPA-T8, # 301041 [1:100]), CD39 (A1, # 328205 [1:100]), CD69 (FN50, # 310911 [1:100]), CD137 (4B4-1, # 309820 [1:200]) all BioLegend, CD25 (2A3, #340939 [1:20]), CD134 (L106, # 340420 [1:10])-BD Biosciences. Samples were acquired on the Aurora CS spectral flow cytometer (Cytek Biosciences, USA) using the Spectroflo software v3.0 (Cytek). Prior to each run, all samples were fixed in 0.5% paraformaldehyde. Data analysis was performed using FlowJo version 10.7.1 (BD Biosciences).

##### Virome capture sequencing

Virome capture sequencing was performed on stool swabs collected at 4 and 8 months suspended in PSP/Enat buffer, using the VirCapSeq-VERT hybrid-capture platform (Roche) as previously described ^20^. Total nucleic acid was extracted using the AllPrep PowerViral DNA/RNA kit (QIAGEN), and RNA was converted to double-stranded cDNA using random hexamers. Sequencing libraries were prepared from approximately 100 ng of nucleic acid using the KAPA HyperPlus kit (Roche) with unique dual-index adapters. Libraries were pooled by equal mass and hybridized for 16 h to VirCapSeq-VERT probes targeting known vertebrate-infecting viruses, following the SeqCap EZ HyperCap workflow and manufacturer’s instructions. Enriched libraries were PCR amplified and sequenced on an Illumina NovaSeq 6000 platform using 1 × 100-bp single-end reads. Reads were analysed using the CZ ID (formerly IDseq ^21^) metagenomic pipeline. Adapter and human host sequences were removed, followed by nucleotide- and protein-level taxonomic classification, de novo assembly and read remapping to viral contigs. Viral detections were defined using the laboratory’s prespecified threshold of at least 100 nucleotide reads matched by BLAST at the species level (based on proximity to targeted qPCR limit of detection defined previously ^22^ with extraction controls included to monitor contamination. All remaining extraction, library preparation, capture and sequencing steps were performed according to the manufacturer’s instructions.

##### CMV and EBV PCR

The presence of CMV and EBV DNA in plasma was assessed by qPCR on CFX96 (Bio-Rad). Total nucleic acid was extracted from plasma using the MagMAX™ Viral/Pathogen Ultra Nucleic Acid Isolation Kit (ThermoFisher Scientific) on a KingFisher Flex automated purification system, according to the manufacturer’s instructions and laboratory SOP. CMV and EBV DNA were detected and quantified using the RealStar CMV PCR Kit 1.0 and RealStar EBV PCR Kit 1.0 (altona Diagnostics GmbH, Hamburg, Germany), according to manufactures instructions. Both assays use virus specific primers and fluorescently labelled probes for real time detection of amplified viral DNA and incorporate a heterologous internal control to monitor assay performance and PCR inhibition. Viral DNA concentrations were determined against four assay specific quantification standards (QS1–QS4; 10,000, 1,000, 100 and 10 IU/µL) to generate a standard curve for quantitative analysis.

##### Live virus neutralization assay

Rapid high-content neutralization assay with HAT-24 cells was done as previously described ^23^. Briefly, human sera were serially diluted (1:2 series starting at 1:10) in DMEM-5% FBS and mixed in duplicate with an equal volume of SARS-CoV-2 virus solution standardized at 2× VE_50_ ^24^. After 1 h of virus–serum coincubation at 37 °C, 40 μL were added to an equal volume of nuclear-stained HAT-24 cells pre-plated in 384-well plates as above. Plates were incubated for 20 h before enumerating nuclear counts with a high-content fluorescence microscopy system as indicated above. The % neutralization was calculated with the formula: %N = (D − (1 − Q)) × 100/D as previously described.^1^ Briefly, “Q” is a well’s nuclei count divided by the average count for uninfected controls (defined as having 100% neutralization) and D = 1 − Q for the average count of positive infection controls (defined as having 0% neutralization). Sigmoidal dose–response curves and NT_50_ values (reciprocal dilution at which 50% neutralization is achieved) were obtained with GraphPad Prism software.

##### Single cell RNA-seq analysis PBMCs and sequence alignment

Briefly, PBMC from 20 individuals from ADAPT cohort (10 LC and 10 MC) at 4-, 8- and 24-month timepoints were genotyped and had PBMCs sequenced using the 10x genomics platform. For each individual, 1 vial (∼1mL) of biobanked (frozen) PBMCs was retrieved and thawed. Cell density per vial was roughly 7×10^6^ cells/mL. To this end ∼30uL was used for single-cell RNA-sequencing, and another fraction (10^6^) for genotyping. Cell viability was tested using trypan blue, with high levels ∼>90% for all samples. Live cells were pooled (multiplexed) with 10 participant samples per pool in replicate, for a total of 4 batches. For each batch, single-cell RNA capture and barcoding with the Single Cell 5’ v2.0 NextGEM single cell RNA seq (GEX (#PN-1000165) + VDJ (TCR+BCR) (#PN-1000005)) from 10x Genomics, with a target capture of 20,000 cells was performed. Sequencing was done with the Illumina NovaSeq 2000 on a S4 flowcell. We aimed for 30,000 Reads Per Cell (RPC) for GEX, and 7,500 RPC for both BCR and TCR respectively. Reads were processed and demultiplexed the using Cell Ranger Single Cell Software Suite (v 7.0.0; 10x Genomics). Mapping and alignment were done to GRCh38 (GEX: refdata-cellranger-GRCh38-2020-A, and VDJ: refdata-cellranger-vdj-GRCh38-alts-ensembl-5.0.0) using STAR within the Cell Ranger Suite. The pipeline was executed on a high-performance cluster with a 3.10.0-1160.42.2.el7.x86_64 operating system.

##### Genotyping and demultiplexing

DNA was extracted using Qiagen QIAamp DNA mini kit and genotyping was done using the UKB Axiom array. The genotypes were called using the Axiom Analysis Suite from Thermofisher (AxAS v5.1) following the Best Practices Genotyping Analysis Workflow in the Axiom_UKB_WCSG.r5 library. Imputation was performed using the Michigan Imputation Server with Eagle (v2.3) for phasing, and Minimac4 (v1.0.2) for imputation and the 1000 Genomes project reference panel (1000G 30X WGS reference panel). Missingness was assessed with vcftools (v0.1.16). Two samples (AD007 and AD322) were rerun with adjusted parameters due to high missingness (>5%). After imputation, we filtered on MAF>0.05 and R2>0.3 using bcftools (v1.10.2.). Demultiplexing was performed to assign cells to individuals using Demuxlet.

##### Quality control and cell classification

Each pool was processed individually, with cells assigned as doublets excluded from the analysis, along with cells with higher than 5% mitochondrial gene expression. Following the quality control processing, Seurat was used to integrate the individual pools after normalisation using the SCTransform function. To cluster and classify cells, VDJ genes were removed from the integrated object counts by matching gene names to immunoglobulins (IG) with: “IG[HLK][VJ]” or “IGHD-”, and T-cell receptors (TR): “TR[ABGD][VDJ]”. SCTransform was then used on “non-VDJ” count data to normalise the expression levels. Cells were then classified using Azimuth/Seurat pipeline with the human PBMC reference for L2 and the metadata was then added to the full count expression data. All further analyses were performed using the integrated object with the full expression data in R Statistical Software (v4.1.3; R Core Team 2022).

##### Single-cell analysis and cell scores

With the full object, cell-type proportions were calculated for individuals split by long-COVID status and tested for significance using the propeller function in the speckle R package. Using the AddModuleScore function in Seurat, a CD8 exhaustion score and an interferon score (IFN) were calculated, along with T stem cell memory and T cytotoxic scores, based on gene sets from Khoo et al19,25.

##### Post-processing of 10x VDJ datasets

Filtered VDJ contigs generated by Cell Ranger were post-processed with IgBLAST (version 1.1.9) to align against the IMGT human Reference Directories to generate AIRR-C tab-delimited output. For cells with multiple chains for the same loci the chain with the highest UMI count was retained. T and B cell VDJs were filtered to remove non-productive chains (stop codons or out-of-frame) and chains that lacked CDR3’s and analysis was restricted to cells that were Singlets.

To defined B cell clonal lineages, CDR3s from B cell IGH/K/L were extracted and CDR3 nucleotide sequences were binned based on V gene, J gene and CDR3 length for clustering with cd-hit (version 4.7) using the cd-hit-est tool. Clustering was undertaken at a 90% identity threshold for cells from each individial and B cells that shared the same cluster membership for IGH and IGK/L were considered clonally related. For T cells, clonotypes were defined by shared V gene, J gene and CDR3 amino acid sequence.

To annotate putative antigen specificity for T cells two databases of TCRs of known specificity were obtained; immuneCODE for SARS-CoV-2 specific TCRs and VDjdb (version 2022-02-30) for SARS-CoV-2 plus other antigens reported in literature. TCR clonotype putative specificity were annotated by exact TRB clonotype matches (same V, J and CDR3 AA sequence for TCR beta loci). Where the same clonotype was associated with more than one antigen the TCR specificity was flagged as ‘multiple’. For the B cells, to explore SARS-CoV-2 specificity, sequences were obtained from CoV-AbDab (version 210223. CDR3 AA sequences from the IGH of known specificity were clustered with CDR3s from the 10x datasets using cd-hit at an 80% identity threshold. If an IGH of known specificity clustered with IGH from 10x then those cells were annotated with the specificity from the CoV-AbDab database. VDJ data from Cell Ranger and IgBLAST were integrated with GEX analysis and putative antigen specificity in R (version 4.3.0) within RStudio IDE (version 2023.3.1.446) using the tidyverse package (version 2.0.0).

##### Statistical Analysis

All graphs are presented as medians with inter-quartile ranges. For unpaired samples Mann-Whitney U test was used, Kruskal-Wallis with Dunn’s correction was used for multiple comparisons, linear mixed-effects models were used to determine temporal changes, all employing Prism 10 (GraphicPad, La Jolla, CA, USA) software. *p* values <0.05 were considered significant (*<0.05, **<0.01 and ***<0.001). Correlograms and PCA analyses were performed on R (version 4.3.0). Analysis of patient reported outcome measures was conducted using Stata v14 (StataCorp LLC, College Station, TX, USA).

## Data availability

Clinical data from this study are held by the data management team, and can be made available to other research groups, after approval of a proposal by ADAPT steering committee. To protect patient privacy, underlying electronic health records may be accessed via a remote server pending data access agreement. Please contact the corresponding author with an outline of the intended use. scRNAseq data generated in this study have been deposited in a public database under accession code SRA: SRP497951, Bio Project: PRJNA1092125 and GEO: GSE262861. All other source data are provided as a Source Data file.

## Code availability

Codes used for scRNAseq analysis have been deposited in the Zenodo database. Access to codes can be found here Doi:10.5281/zenodo.10888516.

## Acknowledgements

We thank the staff at the St Vincent’s Centre for Applied Medical Research-Clinical Trials Unit for their expertise in specimen processing and biobanking. We thank E. Johansson Beves for assistance with the Cytek Aurora, and J Caguicla and Z Naing at POW diagnostic serology lab. We appreciate grant support from the St Vincent’s Clinic Foundation, the Curran Foundation, the Rapid Response Research Fund, and the Medical Research Futures Fund (Australia). CP, RVJ and RMRK was supported by MRFF grants; 2024/MRF2035199, 2025/MRF2047230. S.B. was supported by a fellowship from the Magid’s and a Ramsay award by the Solve ME/CFS Initiative (SMCI).

## Author contributions

Protocol design and clinical management: GVM, GJD, DD, ADK, PC, AB, BJB. Experimental design and procedures: CP, RVJ, RMRK, KLJ, SB, LN, AAg, SGT. Data analysis and Visualization: CP, RVJ, RMRK, NFA, BJ, KLJ, SB. Writing - original draft: CP, RVJ, RMRK, ADK. Writing - review & editing: CP, RVJ, RMRK, KLJ, BJ, ADK, GVM, GJD, BM, VK, KLJ, SB, DBW. All authors contributed to the manuscript and approved the submitted version.

