## Supplementary figures for "Altered herpesvirus-specific immune response is associated with long COVID"

### Supplementary Table 1

Table 1: Characteristics of ADAPT participant at time of enrolment

| | CC | LC | |
|---|---|---|---|
| | 0 (n = 53) | 1 (n = 52) | Total (n = 105) |
| Age | 50 (23, 75) | 50.5 (27, 83) | 50 (23, 83) |
| Sex | | | |
| Female | 27 (50.9%) | 29 (55.8%) | 56 (53.3%) |
| Male | 26 (49.1%) | 23 (44.2%) | 49 (46.7%) |
| Caucasian/white ethnicity | | | |
| No | 7 (13.2%) | 5 (9.6%) | 12 (11.4%) |
| Yes | 46 (86.8%) | 47 (90.4%) | 93 (88.6%) |
| Comorbid medical condition(s) | | | |
| No | 28 (52.8%) | 28 (53.8%) | 56 (53.3%) |
| Yes | 25 (47.2%) | 24 (46.2%) | 49 (46.7%) |
| Body Mass Index | 23.6 (0, 44.7) | 24 (0, 35.9) | 23.7 (0, 44.7) |
| Days since diagnosis | 124 (88, 364) | 125.5 (97, 242) | 124 (88, 364) |
| Enrolment source | | | |
| Community | 50 (94.3%) | 39 (75%) | 89 (84.8%) |
| Hospital inpatient | 3 (5.7%) | 13 (25%) | 16 (15.2) |
| Symptom severity at initial infection | | | |
| Mild | 39 (73.6%) | 1 (1.9%) | 40 (38.1%) |
| Moderate | 14 (26.4%) | 51 (98.1%) | 65 (61.9%) |
| Symptoms at enrolment visit | | | |
| Shortness of breath | 1 (1.9%) | 24 (46.2%) | 25 (23.8%) |
| Chest pain | 0 (0.0%) | 14 (26.9%) | 14 (13.3%) |
| Diarrhoea | 1 (1.9%) | 8 (15.4%) | 9 (8.6%) |
| Malaise/weakness | 0 (0.0%) | 16 (30.8%) | 16 (15.2%) |
| Fatigue | 4 (7.5%) | 40 (76.9%) | 44 (41.9%) |

### Supplementary Figure S1

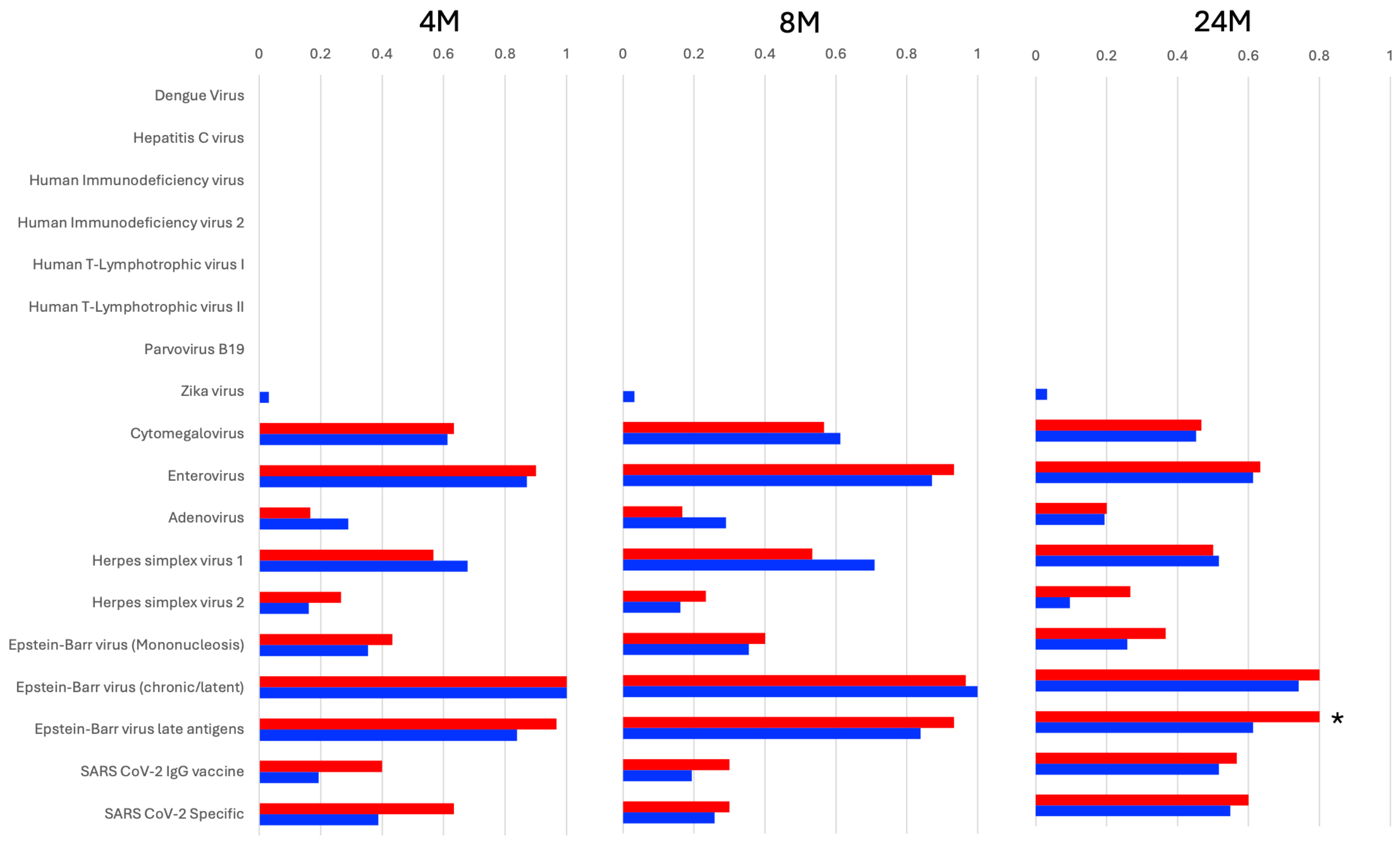


**Figure S1: Seropositivity across viral antigen groups over time.** Proportion of participants with long COVID (LC; red) and convalescent controls (blue) who were seropositive for the indicated viral antigen groups at 4, 8 and 24 months after infection. The asterisk indicates the nominal difference in seropositivity to EBV late antigens at 24 months. Other comparisons were not significant.

### Supplementary Figure S2

A

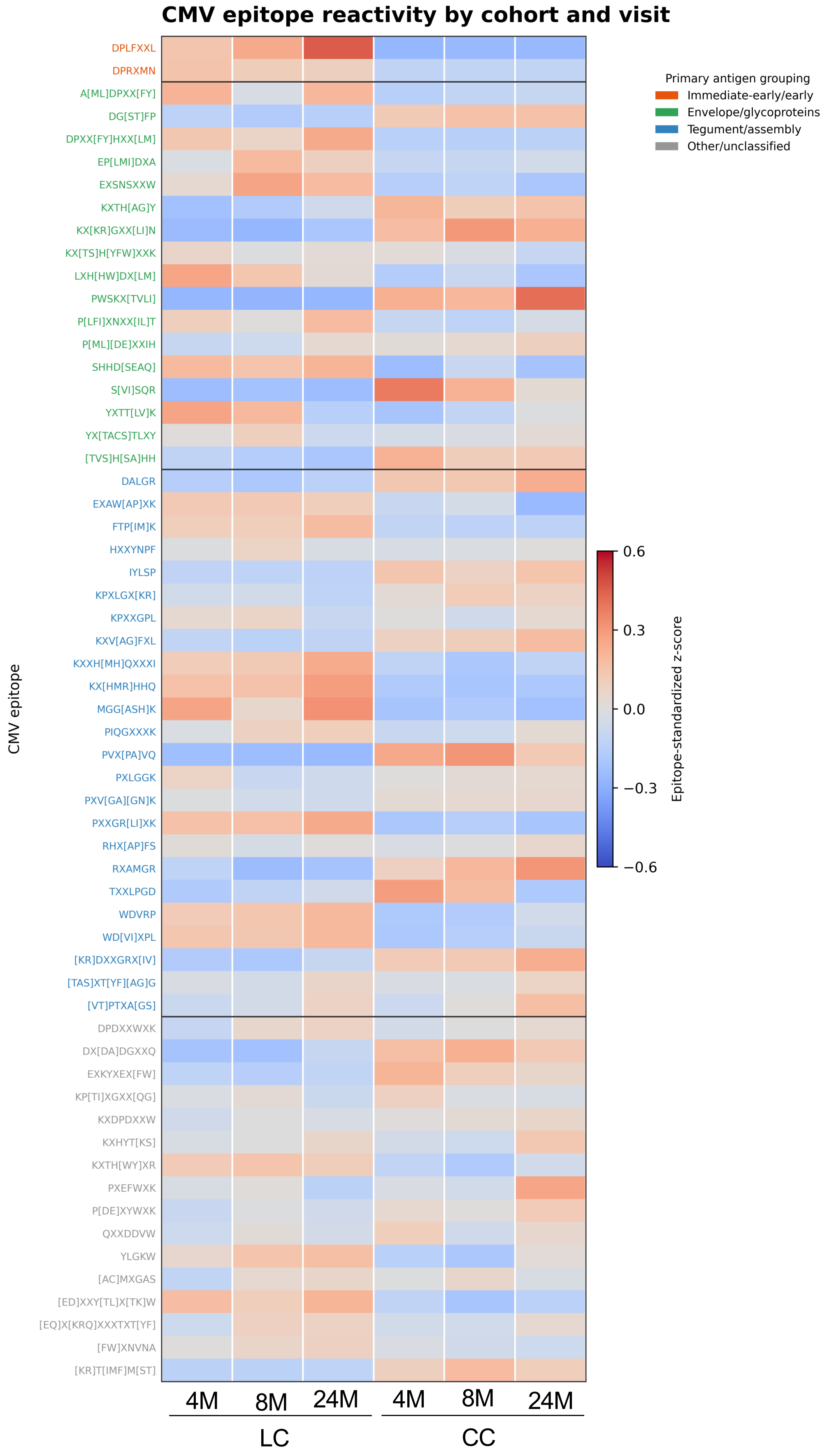

Figure S2

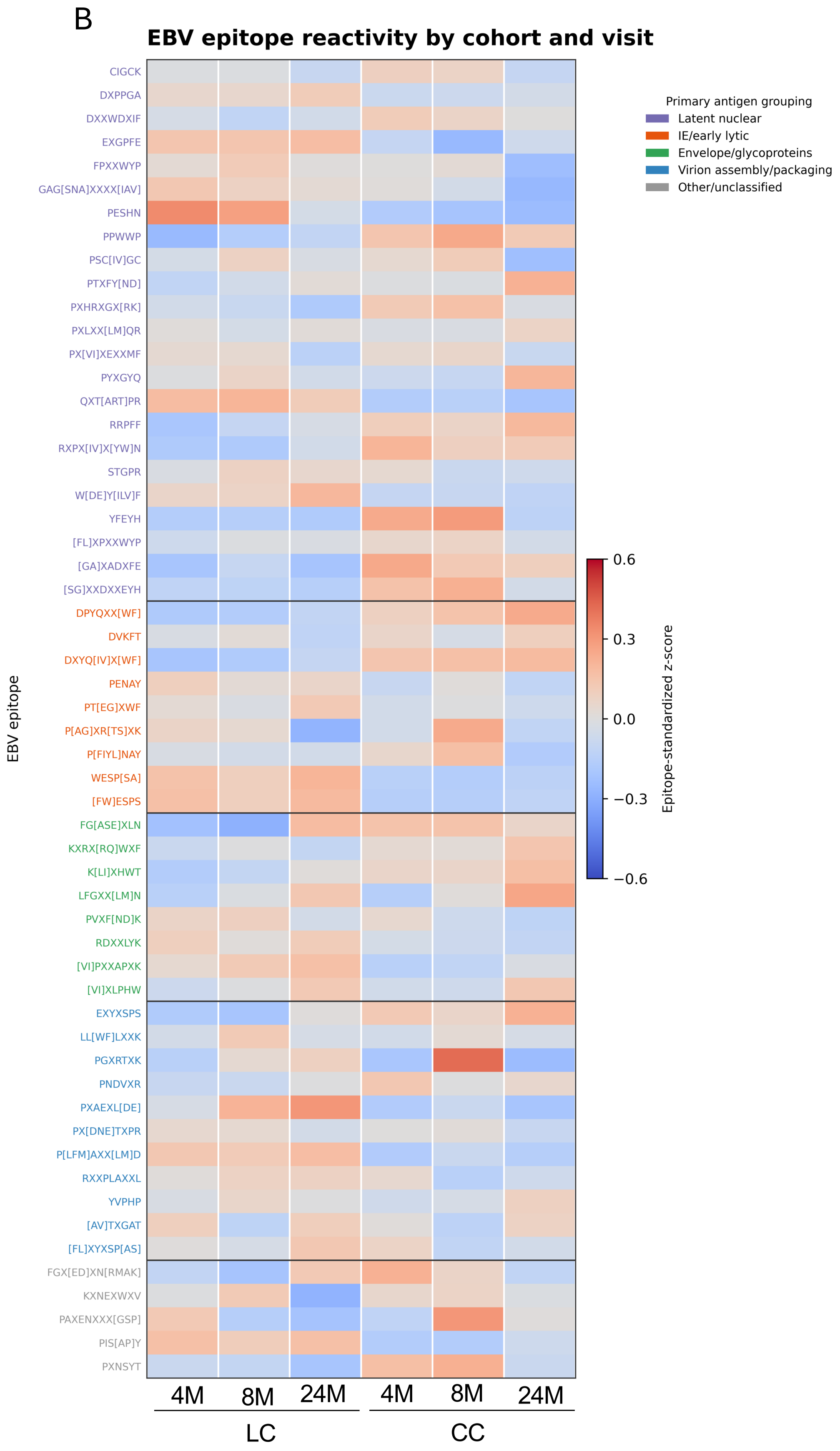


**Figure S2: CMV- and EBV-specific epitope reactivity by cohort and visit.** Heatmaps show mean epitope-standardized z-scores for (A) CMV and (B) EBV epitopes in LC and matched controls at 4, 8 and 24 months. Rows represent individual epitopes and columns represent visit and groups. Red and blue indicate higher and lower mean z-scores, respectively, as shown by the colour scale. Epitope labels are coloured according to primary antigen grouping: immediate-early/early, envelope/glycoproteins, tegument/assembly and other/unclassified for CMV; latent nuclear, early/lytic, envelope/glycoproteins, virion assembly/packaging and other/unclassified for EBV. Values are means across participants within each cohort and visit.

### Figure S3

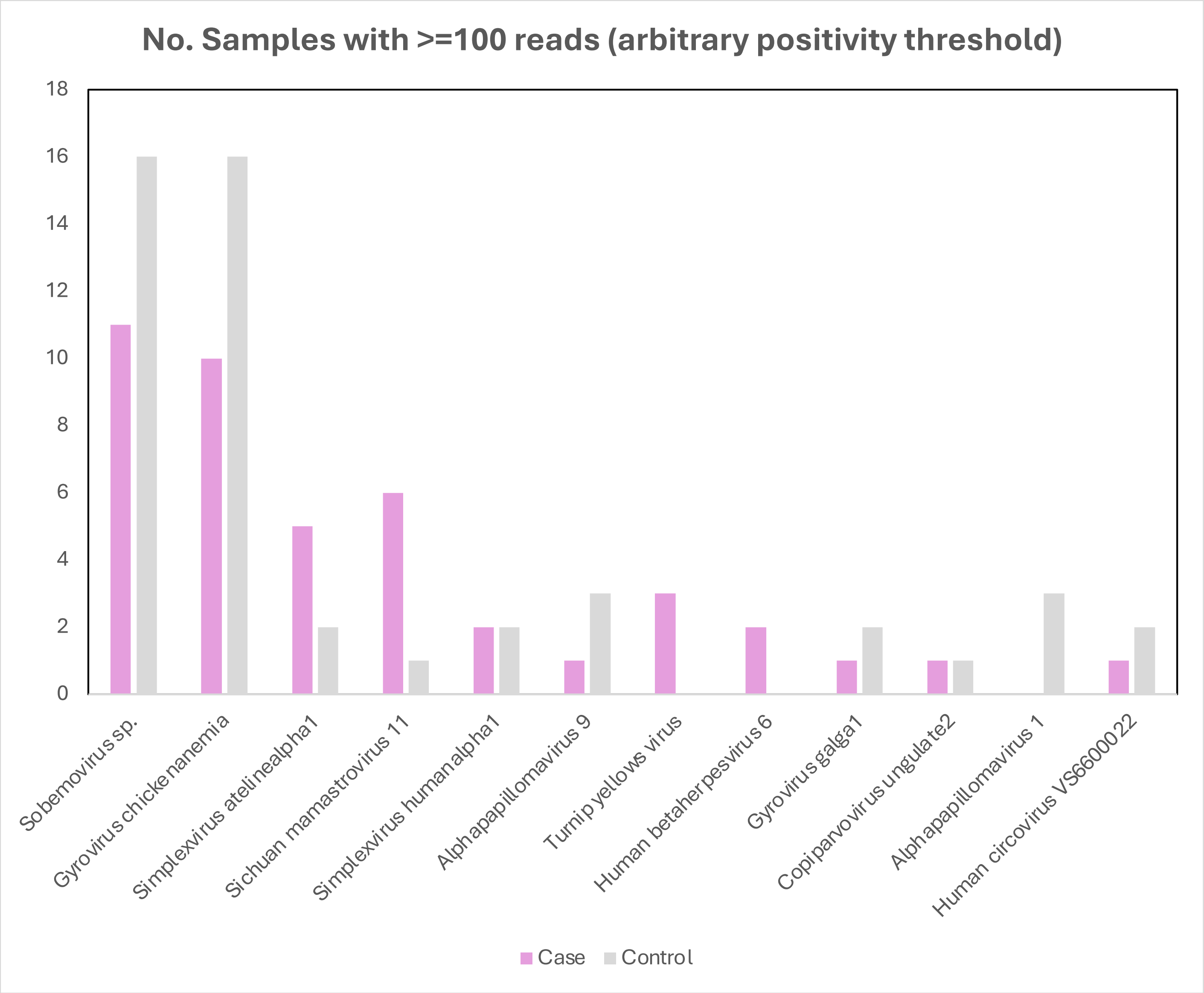


**Figure S3: Viral taxa detected in stool samples by virome capture sequencing.** Bars show the number of LC ("Case"; pink) and convalescent control (grey) samples with at least 100 species-level BLAST-matched reads for each indicated viral taxon. The plot includes taxa detected in more than three samples above this positivity threshold. The threshold was exploratory; no statistical comparisons are shown.

### Figure S4

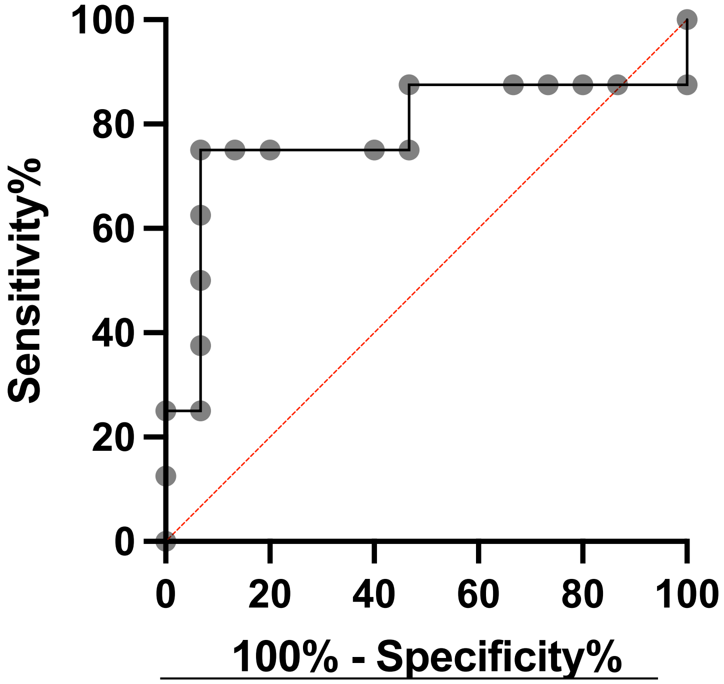


**Figure S4: Receiver operating characteristic analysis of EBV avidity at 24 months.** ROC curve for EBV avidity distinguishing recovered controls from unrecovered LC participants, with recovery defined by EQ-index status at 18–24 months. The analysis included 15 recovered controls and 8 unrecovered LC participants. The AUC was 0.783 (95% CI, 0.541–1.000; *P*=0.0282). The diagonal line indicates chance-level discrimination. AUC, area under the curve; ROC, receiver operating characteristic.
